# GABA_A_ receptor availability in clinical high-risk and first-episode psychosis: a [^11^C]Ro15-4513 positron emission tomography study

**DOI:** 10.1101/2025.02.27.25322861

**Authors:** Paulina B Lukow, Julia J Schubert, Mario Severino, Samuel R Knight, Amanda Kiemes, Nicholas R Livingston, James Davies, Andrea de Micheli, Thomas J Spencer, Paolo Fusar-Poli, Beata Haege, Natasha Vorontsova, Jacek Donocik, Eugenii A Rabiner, Anthony A Grace, Steven C Williams, Philip McGuire, Mattia Veronese, Federico E Turkheimer, Gemma Modinos

## Abstract

Disrupted gamma-aminobutyric acid (GABA) neurotransmission may contribute to the pathophysiology of schizophrenia. Reductions in hippocampal GABAergic neurons have been found in schizophrenia, and increased hippocampal perfusion has been described in schizophrenia and in people at clinical high-risk for psychosis (CHRp). We have also found decreases in hippocampal GABA_A_ receptors containing the α5 subunit (GABA_A_Rα5) in a well-validated neurodevelopmental rat model of relevance for schizophrenia. Positive allosteric modulation of these receptors in the hippocampus using a specific compound was shown to reverse the behavioural and neurophysiological phenotypes of this model. However, whether GABA_A_Rα5 availability is dysregulated in the psychosis spectrum at the regional or network levels is unknown. We addressed this issue by using [^11^C]Ro15-4513 and positron emission tomography (PET) in 22 individuals at CHRp, 10 people with a first-episode psychosis (FEP) and 23 healthy controls (HC). We quantified GABA_A_Rα5 availability in the hippocampus and across the brain, and employed a perturbation covariance method to assess individual molecular covariance deviations in CHRp and FEP groups compared to the HC group. Hippocampal GABA_A_Rα5 availability was not significantly different between groups (*F*(2,50)=0.25, p=0.78). However, network analysis identified significant deviations in GABA_A_Rα5 covariance between groups, both across all regions (all p<0.001, pairwise Cohen’s d = 0.07-0.5) and relative to the hippocampus (all p<0.001, pairwise Cohen’s d = 0.01-0.67). These findings suggest that psychosis symptoms are associated with alterations to the brain-wide organisation of the GABA_A_Rα5 system, rather than changes at a regional level.

## Introduction

There is growing interest in the role of gamma-aminobutyric acidergic (GABAergic) dysfunction in psychosis. The loss of GABAergic interneurons in the hippocampus has been found in post-mortem studies of people with schizophrenia [1–3] and increased hippocampal perfusion has been found in individuals with schizophrenia *in vivo* [4,5]. Similar findings have been reported in people at clinical high-risk for psychosis (CHRp) [6–8], who experience sub-threshold psychotic symptoms [9–11] and are at 25% risk of transition to frank psychosis [12]. Despite extensive research, the biological mechanisms underlying different degrees of psychosis symptoms remain largely unknown [6–8].

A mechanistic investigation of hippocampal hyperactivity in psychosis development has been provided by studies employing the methylazoxymethanol acetate (MAM) rodent model. This model involves administration of the mitotoxin MAM to pregnant rats on gestational day 17, which interferes with hippocampus development [13] and results in local hyperactivity [14]. Upon reaching adulthood, MAM rats present with a number of behaviours consistent with a psychosis-relevant phenotype: elevated response to amphetamine administration, a decrease in social interaction, reduced executive function, a lower startle threshold and a deficit in attenuating startle response to repetitive stimuli [13]. Additionally, they exhibit elevated dopamine release into the striatum [14,15], a phenomenon found robustly in psychosis and schizophrenia [16] and two independent samples of CHRp individuals [17]. The hippocampus regulates striatal function through direct projections [18] and indirectly through ventral tegmental area (VTA) disinhibition [19,20]. Interestingly, MAM rats also show alterations to the GABAergic system in the hippocampus, providing insight into the molecular mechanisms linking hippocampal dysfunction, striatal hyperdopaminergia and psychosis.

GABAergic interneuron loss has been found in the hippocampus of adult MAM rats [21]. This finding is analogous to the generally decreased nonpyramidal cell numbers [1], and a reduction in specific cell-type densities [2] and numbers [3] in the hippocampus in schizophrenia. Interestingly, peripubertal amplification of GABAergic function in MAM rats with the benzodiazepine diazepam was shown to prevent MAM phenotype emergence and GABAergic inhibitory interneuron loss in the hippocampus [21,22]. However, benzodiazepines show affinity for several GABA_A_ receptors expressed across the brain [22]. In this context, GABA_A_ receptors containing the α5 subunit (GABA_A_Rα5) are an attractive target due to being predominantly expressed in the hippocampus [23]. Direct infusion into the hippocampus of a selective GABA_A_Rα5 positive allosteric modulator in adult MAM rats was shown to normalise the dopaminergic output from the VTA into the striatum, and the exaggerated hyperlocomotor response to amphetamine [24]. Furthermore, genetic α5 subunit deletion in mice was found to produce a phenotype similar to that of adult MAM rats [24]. Moreover, our recent *ex vivo* autoradiography study with [^3^H]Ro15-4513 in MAM rats showed lower density of GABA_A_Rα5 across the hippocampus [25]. Altogether, this suggests a potential role of GABA_A_Rα5 alterations in psychosis, making them a possible novel therapeutic target for this disorder.

GABA_A_Rα5 represent about 30% of all GABA_A_Rs in the hippocampus, but only 7-8% of them across the brain [26]. They are predominantly extrasynaptic and show high affinity for GABA, which facilitates tonic inhibition of neural firing in response to ambient neurotransmitter levels [27,28]. A reduction in GABA_A_Rα5-mediated tonic inhibition in the hippocampus could lead to an increase in local pyramidal neuron firing rate, resulting in hyperactivity. However, evidence supporting such hypothesis from human studies remains limited. Very few post-mortem studies assessed hippocampal GABAergic receptor availability in schizophrenia, and neither used a label selective for receptors containing a specific subunit [29–31]. Post-mortem studies may be affected by long illness duration, medication exposure and death, further limiting interpretability. Here, positron emission tomography (PET) can be used to measure receptor availability in the human brain *in vivo* in different populations from the psychosis spectrum.

The two most common GABA_A_R PET radiotracers are radiolabelled flumazenil and Ro15-4513 [32]. While the former has similar affinity for a number of GABA_A_Rs [33], the latter shows high specificity for GABA_A_Rα5, to which it binds allosterically [34]. To date, GABA PET studies in people with different degrees of psychosis symptoms using either radiotracer have been inconclusive. Experiments in schizophrenia using radiolabelled flumazenil found inconsistent results, with only one [35] of the two [35,36] available studies reporting increased binding in the hippocampus of antipsychotic-naïve schizophrenia patients. Studies employing [^11^C]Ro15-4513 found unchanged [37] or decreased [38] binding in the hippocampus in schizophrenia. The only GABA PET study in the CHRp population found no changes in [^18^F]flumazenil binding in the hippocampus [39]. However, several of these studies employed quantification relying on comparison to a reference region, an approach which may have not been sensitive enough for the study of GABAergic function, as no region in the brain is devoid of GABA_A_Rs [40]. Therefore, an investigation of GABA_A_Rα5 availability in individuals at CHRp and with early psychosis using full tracer kinetic quantification is warranted.

While evidence suggests that neurotransmission may be altered regionally in psychosis, it has also been posited that the symptomology may emerge from network-level dysfunction [41]. A novel method that allows addressing this question using neuroimaging data is the perturbation network covariance approach [42,43]. This is a statistical framework used to construct single-participant-specific networks that reveal abnormalities in e.g., individual molecular covariance patterns relative to a reference sample (e.g., healthy controls) [44]. These patterns are then averaged across all participants to investigate group perturbations across the network. We sought to complement standard group comparisons by additionally employing the kinetic modelling quantification parameters in this network-based approach, to comprehensively characterise brain-wide GABA_A_Rα5 organisation in people with different degrees of psychosis symptoms.

The present study aimed to use [^11^C]Ro15-4513 PET with full tracer kinetic quantification methods in CHRp individuals and people with a first-episode of psychosis (FEP) to examine regional binding; and, to conduct a brain-wide perturbation covariance analysis to evaluate individual molecular covariance deviations from the reference healthy controls. We hypothesised that there would be a reduction in [^11^C]Ro15-4513 binding in the hippocampus of individuals at CHRp and people with an FEP; and that both CHRp and FEP individuals would exhibit significant molecular inter-regional deviations relative to the healthy control group.

## Methods

### 1. Participants

Twenty-four individuals at CHRp and ten with an FEP were recruited from the South London and Maudsley NHS Trust [45]. Twenty-four healthy controls (HC) were additionally recruited through public advertisement from the same geographical area. All participants had capacity for consent, were between 18-40 years old, had no neurological or major medical conditions (e.g., aneurysm, severe asthma), had an IQ≥70 estimated by the Wechsler Adult Intelligence Scale (WAIS-III) [46], presented adequate collateral circulation in the hand, had no history of clotting or renal abnormality, were not currently exposed to any drug with a GABAergic or glutamatergic mechanism of action, had no contraindication to MRI, did not donate >500ml of blood within 60 days prior to scanning, did not have radiation exposure exceeding >10mSv in preceding 12 months, had no past or present history of substance abuse, and were not pregnant or breastfeeding. Healthy blood clotting was ascertained with the international normalised ratio test and the activated partial thromboplastin time test. CHRp state was defined by presence of current attenuated psychosis symptoms, according to the Comprehensive Assessment for At-Risk Mental States (CAARMS) [47]; participants with additional brief limited intermittent psychotic symptoms (BLIPS) [48–51] or family history of psychotic disorder were included. Only CHRp participants without any previous or current history of psychosis (excluding BLIPS) or antipsychotic exposure were included. CHRp participants with current or past antidepressant exposure were included. Additional inclusion criteria for FEP participants included an ICD-10 diagnosis of psychotic disorder (F20-F29 and F31) [52], at least one rating of moderate severity on the positive subscale of the Positive and Negative Syndrome Scale (PANSS) [53] and ‘first treatment contact’, i.e., receiving support from an Early Intervention mental health service for a period of less than two years since first admission. Participants with an FEP were included if they never received any antipsychotic medication, were not currently taking antipsychotic medication, or were on a stable dose of any antipsychotic other than clozapine and were not treatment refractory. Additional inclusion criteria for HC participants were no self-reported history of Axis I mental health disorder and no personal or familial history of psychosis. The study obtained ethical approval from the London/Surrey Research Ethics Committee (17/LO/1130). All participants provided written informed consent before participation, in accordance with The Declaration of Helsinki [54].

### 2. PET acquisition

Scanning was performed on a SignaTM PET-MR General Electric (3T) scanner using the MP26 software (01 and 02) at Invicro, an imaging centre located in London, UK. The radiotracer was administered through the dominant antecubital fossa vein in a single bolus injection, administered at the beginning of the scanning session. Arterial blood sampling was performed from the radial artery in the non-dominant wrist for the production of a metabolite-corrected plasma input function. Continuous blood sampling was performed during the first 15 minutes of the scan, followed by up to 13 discrete samples (maximum 195ml in total, including samples required for arterial line clearing). The mean ± SD amount of radiation administered was 283.25 MBq ± 72.79 (range: 124.13– 404.75 MBq). PET acquisition was performed in 3D list mode for 70 minutes and binned in the following frames: 15sx10, 60sx3, 120sx5, 300sx11. Attenuation correction was performed with a ZTE sequence (voxel size: 2.4 × 2.4 × 2.4 mm^3^, field of view = 26.4, 116 slices, TR = 400 ms, TE = 0.016 ms, flip angle = 0.8°). A T1-weighted IR-FSPGR sequence was used for PET image co-registration (voxel size: 1 × 1 × 1 mm^3^, field of view = 25.6, 200 slices, TR = 6.992 ms, TE = 2.996 ms, TI = 400 ms, flip angle = 11°). Image reconstruction was done by ordered subset expectation maximization reconstruction with point spread function recovery and time-of-flight activated. A power calculation using G*Power on [^11^C]Ro15-4513 PET data from a previous study [55] indicated that 20 participants per group were required to detect a significant change in binding with a power of 0.8 (alpha = 0.05, two-tailed).

### 3. Demographic and clinical data analysis

Clinical information about the study sample included psychosis symptom severity (assessed with CAARMS in CHRp and HC, and with PANSS in FEP), anxiety and depression symptom severity (Hamilton Anxiety [56] and Depression [57] Rating Scales), current functioning (Global Functioning: Social and Role Scales [58]) and current neuroleptic medication. Demographic information included age, sex and ethnicity. Chlorpromazine equivalent doses of antipsychotic medication for people with an FEP on a stable dose were calculated using the *chlorpromazineR* package (https://docs.ropensci.org/chlorpromazineR/) in R 4.2.1 using the previously published conversion method [59–63].

Analysis of clinical and demographic data was performed in R 4.2.1. χ^2^ test was used to analyse differences in sex between groups with the *chisq.test()* function. One-way ANOVA with the *anova_test()* function from the *rstatix* package (https://CRAN.R-project.org/package=rstatix) was used to compare differences between groups in other demographic and clinical variables. Post-hoc pairwise t-tests were used to determine differences between groups with the *pairwise_t_test()* function from the *rstatix* package, with the Holm multiple comparison correction method [64]. Results in all analyses were considered significant at p<0.05. Missing data were omitted from analysis.

### 4. PET data quantification and quality checks

PET images were pre-processed with MIAKAT 4.3.24 in MATLAB R2017a. For each participant, an isotropic, skull-stripped IR-FSPGR structural image linearly transformed to the MNI template [65] was co-registered to an isotropic, motion-corrected integral image created from the PET time series. The CIC v2.0 neuroanatomical atlas [66] was non-linearly transformed to the participant’s structural image and used to define the following regions of interest (ROIs): whole-brain, whole-brain grey matter, cortex, subcortical regions, cerebellum, brain stem, occipital lobe, insular cortex, temporal lobe, frontal lobe, cingulate cortex, parietal lobe, basal ganglia, thalamus, parahippocampal gyrus, amygdala, hippocampus, anterior cingulate cortex, globus pallidus, striatum, caudate, nucleus accumbens and putamen. Time-activity curves were extracted for each region of interest individually from the dynamic PET acquisition using the CIC v2.0 atlas [66] and then input into a two-tissue compartmental model using the regional brain tissue and the blood as the two compartments. The outcome measure from the two-tissue compartmental model was the total volume of distribution (V_T_), in agreement with Horder et al. [67]. Pre-processing outputs (brain segmentation, motion correction, left-right image orientation consistency, motion correction, ROI fitting to individual brains, time-activity curves and parent plasma fraction plots for the hippocampus and whole-brain) were quality-checked visually for each participant. Poor quality data insufficient for quantification or with abnormal model fitting was excluded. Then, model parameters (V_T_; rate constant for transfer from arterial plasma to tissue, K1; fractional volume of blood within a given region, Vb [68]) were assessed for the existence of outliers. A participant was considered for exclusion if at least 50% of their values across all ROIs exceeded the outlier detection threshold, which was at mean +/- 3 standard deviations for the whole study sample for a given ROI. No participants were excluded on this basis. If individual data points within included participant datasets were identified as outliers, they were considered for exclusion if the corresponding time-activity curve did not pass the quality check. No data points were excluded on this basis.

### 5. PET data analysis

Our primary outcome measure was hippocampal [^11^C]Ro15-4513 binding. Between-group comparison of hippocampal [^11^C]Ro15-4513 V_T_ values was performed including the three groups with a one-way ANOVA with age and sex as covariates of no interest. In a supplementary analysis to assess between-group differences in [^11^C]Ro15-4513 binding across the brain, mixed ANOVAs were conducted including all regions tested, with age (mean-centred) and sex as covariates of no interest. All ANOVA models were fitted with the *anova_test()* function from the *rstatix* package. Effect sizes are reported in generalised eta squared (ges) from the *anova_test()* output. Post-hoc pairwise comparisons were performed with the *anova_test()* function from the *rstatix* package for each group pair. Results were considered significant if they passed Bonferroni correction for multiple comparisons. Correlations with symptoms were estimated using the *cor.test()* function from the *stats* package, omitting missing values.

### 6. Covariance perturbation analysis

Covariance perturbation analysis [44] was performed to investigate group differences in brain-wide correlations in [^11^C]Ro15-4513 binding. First, a reference network was constructed using HC data, by performing partial Pearson correlation analysis of [^11^C]Ro15-4513 V_T_ values between each region pair across the HC group, with age and sex included as covariates. Next, a participant-level measure of difference was calculated for each non-HC participant. This was done by adding a single participant from the non-HC group to the HC reference group, and constructing another network as above. This new network was termed the “perturbed” network, in accordance with the nomenclature established by Liu et al. [44]. The difference between the perturbed network and the reference network was then calculated to obtain a differential network measure, denoted as *ΔPCC*. The null hypothesis was that *ΔPCC_n_* is equal to the population mean of *ΔPCC_n_*, and thus:

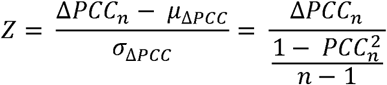

Where n denotes the total number of participants in the reference group, while *µ*_Δ*PCC*_ and σ_Δ*PCC*_ are the mean and standard deviation of the differential network *ΔPCC*. It can be shown that for sufficiently large n (previously indicated as at least 28 participants [69]), the mean and the standard deviation of *ΔPCC_n_* for the population are µ_Δ*PCC*_ =0 and 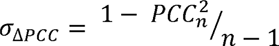 [44]. Thus, the *Z* matrix essentially represents the level of abnormality in the covariance, where each edge exhibits a level of variation in molecular availability, leading to deviations from the values observed in the reference group. The p-value for each edge can be derived from the z-scores [44]. For the HC group, participant-level connectivity matrix was calculated by removing one HC participant at a time (i.e., by jackknife resampling), as done previously [70]. Data points identified as outliers (see ‘PET data quantification and quality checks’ above) were imputed using the mean of the ROI values across all other participants [71]. Values were imputed for three HC (for parahippocampal gyrus, nucleus accumbens and anterior cingulate, respectively) and for one CHRp participant (nucleus accumbens).

### 7. Between-group differences in regional [^11^C]Ro15-4513 binding covariance

Statistical comparisons of the distribution of these deviation measures (z-scores) were conducted using a 1 000-permutation test, wherein the null distribution was estimated by permuting group labels. The magnitude of the effect was quantified using Cohen’s *d* effect size, appropriate for pairwise comparisons. Lower triangular matrices across groups were used for this analysis to exclude duplicate z-score values. These differences were first evaluated across all brain regions, and subsequently only for correlations between the hippocampus and other brain regions. Additionally, the proportion of extreme deviations at the region level was computed for each participant, a metric indicating deviation patterns at the regional level, and statistically compared between groups using a 1 000-permutation test. As the z-score measures and the regional extreme deviations produced similar statistical results, the analysis using z-scores is reported in the main text, while the other is presented in the Supplement for completeness (Supplementary Methods and Results: Regional extreme deviations analysis, Supplementary Figure 3). Additionally, group classification based on covariance networks was performed with a Cubic Support Vector Machine model; this is also reported in the Supplement (Supplementary Methods and Results: Group classification by regional [^11^C]Ro15-4513 binding covariance, Supplementary Figure 4).

## Results

### 1. Demographic and clinical data

Three participants were excluded: one CHRp participant due to missing arterial blood data, one further CHRp participant due to incomplete PET acquisition, and one HC participant due to an error in blood data processing. The resulting final study sample was n(CHRp) = 22, n(HC) = 23, n(FEP) = 10. There was no significant difference in sex between groups (χ^2^=4.2, p=0.12). There was a significant difference in age between groups (*F*(2,52)=3.3, p=0.046, ges=0.11), in that FEP participants were older than the other two groups; however, the results did not survive multiple comparison correction (Supplementary Table 1). There was also a significant difference in reported anxiety and depression between groups (anxiety: *F*(2,50)=19.4, p<0.001, ges=0.44; depression: *F*(2,50)=24.7, p<0.001, ges=0.50), where CHRp participants reported more symptoms of anxiety and depression than FEP and HC participants, and FEP participants reported more symptoms of anxiety and depression than HC (Supplementary Table 1). There was also a significant difference in reported social and role functioning between groups (GF:S: *F*(2,50)=8.7, p<0.001, ges=0.26; GF:R: *F*(2,50)=10.8, p<0.001, ges=0.30), where CHRp and FEP participants reported worse functioning than HC on both subscales (Supplementary Table 1). Participant’s demographic and clinical data is summarised in Table 1.

**Table 1.** Demographic and clinical characteristics of the study sample. ^1^CHRp, clinical high risk for psychosis; ^2^FEP, first episode of psychosis; ^3^SD, standard deviation; ^4^CAARMS, Comprehensive Assessment for At-Risk Mental States (positive items: 1.1.Unusual Thought Content, 1.2.Non-Bizarre Ideas, 1.3.Perceptual Abnormalities, 1.4.Disorganised Speech; negative items: 4.1.Alogia, 4.2.Avolition/Apathy, 4.3.Anhedonia); ^5^PANSS, Positive and Negative Syndrome Scale (positive items: P1-P7; negative items: N1-N7; general items: G1-G16; composite score: negative score subtracted from the positive score); ^6^HAM-A/D, Hamilton Anxiety/Depression Scale; ^7^AP, antipsychotic; ^8^CPZ, chlorpromazine; ^9^GF:S/R Global Functioning Social/Role Scale. GF:S and GF:R scores were missing for two control participants. HAM-A and HAM-D scores were missing for two control participants. Negative CAARMS scores were missing for 10 control participants. Antipsychotic medication dose presented as chlorpromazine equivalents for the seven FEP participants who were taking medication at the time of study. Missing data points were omitted in summary or statistics.

|  | Control<br>(n=23) | CHRp<sup>1</sup><br>(n=22) | FEP<sup>2</sup><br>(n=10) |  |  |
| --- | --- | --- | --- | --- | --- |
| | n | | | $\chi^2/F$ | p |
| Female/Male | 14/9 | 15/7 | 3/7 | 4.2 | 0.120 |
| Ethnicity |  |  |  |  |  |
| White | 7 | 13 | 3 | - | - |
| Black | 2 | 5 | 5 | - | - |
| Asian | 10 | 0 | 0 | - | - |
| Mixed or multiple | 2 | 3 | 2 | - | - |
| Other | 2 | 1 | 0 | - | - |
|  | mean (SD<sup>3</sup>) |  |  | F | p |
| Age | 25.1 (4.5) | 25.1 (4.6) | 29 (3.6) | 3.3 | 0.046 |
| CAARMS<sup>4</sup> |  |  |  |  |  |
| Positive | 2 (2.1) | 12.7 (2.7) | - | - | - |
| Negative | 0.8 (1.5) | 7.1 (4.4) | - | - | - |
| PANSS<sup>5</sup> |  |  |  |  |  |
| Positive | - | - | 17.5 (2.6) | - | - |
| Negative | - | - | 14.9 (6) | - | - |
| Composite | - | - | 2.6 (6.6) | - | - |
| General | - | - | 27.8 (3.4) | - | - |
| HAM-A<sup>6</sup> | 3.8 (2.7) | 17.6 (10.1) | 10.8 (6.3) | 19.4 | <0.001 |
| HAM-D<sup>6</sup> | 2.4 (1.8) | 12.5 (6.4) | 8.8 (4.8) | 24.7 | <0.001 |
| AP<sup>7</sup> (CPZ<sup>8</sup>, mg) | - | - | 204.3 (141.9) | - | - |
| GF:S<sup>9</sup> | 8.2 (1.2) | 6.7 (1.6) | 6.2 (1.6) | 8.7 | <0.001 |
| GF:R<sup>9</sup> | 8.4 (1.1) | 6.9 (1.8) | 6.1 (1.4) | 10.8 | <0.001 |

### 2. Hippocampal [^11^C]Ro15-4513 binding

There was no significant difference in hippocampal [^11^C]Ro15-4513 binding between groups (one-way ANOVA with sex and age as covariates of no interest: *F*(2,50)=0.25, p=0.78, ges=0.01) (Figure 1) or between the CHRp and HC groups, specifically (see Supplementary Results: Hippocampal [^11^C]Ro15-4513 binding).

**Figure 1.**
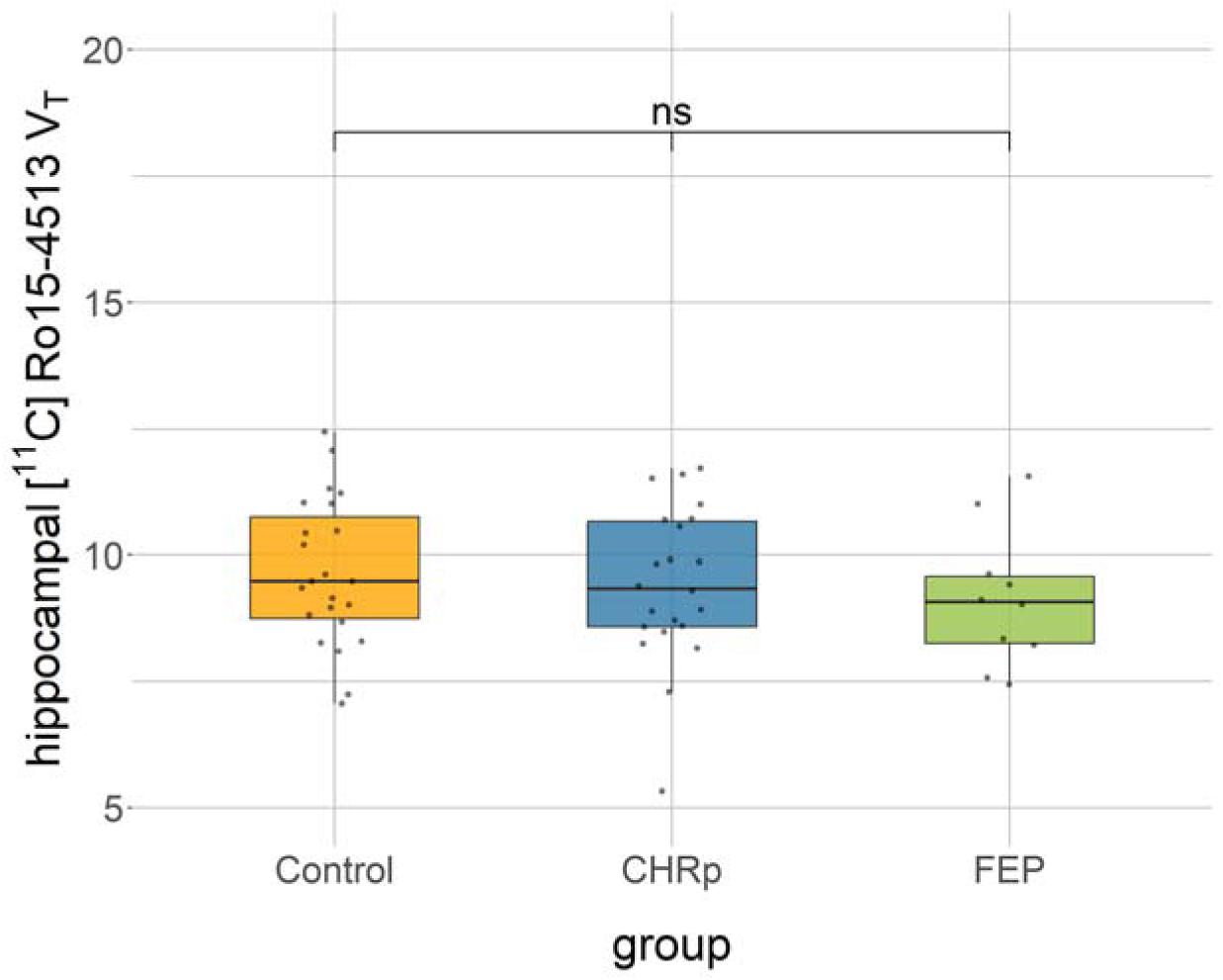
[^11^C]Ro15-4513 binding in the hippocampus between groups. CHRp, clinical high risk for psychosis; FEP, first episode of psychosis; ns, not significant; V_T_, total volume of distribution.

Hippocampal [^11^C]Ro15-4513 binding was not correlated with positive and negative psychosis symptoms or anxiety in the CHRp group. In the FEP group, it was correlated with the severity of negative symptoms, but not of other symptoms. In HC, binding was correlated with levels of positive and negative psychosis symptoms and anxiety. The findings are comprehensively reported in Supplementary Results: Hippocampal [^11^C]Ro15-4513 binding.

### 3. Brain-wide [^11^C]Ro15-4513 binding

A supplementary analysis of [^11^C]Ro15-4513 binding between study groups across all regions tested showed a significant difference between groups (mixed ANOVA with sex and age as covariates of no interest: *F*(2,50)=3.5, p=0.039, ges=0.006) and a significant group*region interaction (*F*(44,1100)=2.3, p<0.001, ges=0.08) (for average binding estimates, see Supplementary Table 2). Pairwise analyses across regions revealed a significant difference between the HC and FEP groups (main effect of group: *F*(1,29)=7.8, p<0.001, p_corr_<0.001, ges=0.01) that was region-dependent (group*region interaction: *F*(22,638)=5.2, p<0.001, p_corr_<0.001, ges=0.15). Post-hoc pairwise comparisons revealed this was driven by lower [^11^C]Ro15-4513 binding in the nucleus accumbens in FEP participants than in the HC group (*F*(1,29)=6.0, p=0.02, ges=0.17), although this result did not survive Bonferroni correction (p_corr_=0.46) (Supplementary Table 3; Supplementary Figure 1).

There was no significant difference between the CHRp and FEP groups across all regions tested (main effect of group: *F*(1,28)=0.006, p=0.94, ges=0.00009). There was a significant group*region interaction (*F*(22,616)=1.9, p=0.007, p_corr_=0.021, ges=0.04), but pairwise post-hoc comparisons did not reveal any region-specific significant differences between the two groups (Supplementary Table 4).

There was no significant difference in [^11^C]Ro15-4513 binding between the HC and CHRp groups across all regions (main effect of group: *F*(1,41)=2.1, p=0.16, ges=0.002; group*region interaction: *F*(22,902)=1.0, p=0.44, ges=0.023).

### 4. Between-group differences in [^11^C]Ro15-4513 binding network covariance

1 000-permutation tests showed significant differences in z-score distributions across all brain regions between HC and FEP (p < 0.001, Cohen’s *d* = 0.5), between HC and CHRp (p < 0.001, Cohen’s *d* = 0.2), but not between FEP and CHRp (p = 0.7, Cohen’s *d* = 0.06). Similar differences were observed when focusing exclusively on the hippocampus, between HC and FEP (p < 0.001, Cohen’s *d* = 0.67), and HC and CHRp (p < 0.001, Cohen’s *d* = 0.26), but not between FEP and CHRp (p = 0.34, Cohen’s *d* = 0.07) (Figure 2). Average z-score matrices across participants in the CHRp and FEP groups can be found in Supplementary Figure 2. These results indicate that the [^11^C]Ro15-4513 binding covariance perturbation network of CHRp individuals exhibited significant negative deviations from HC at a whole-brain level, though to a lesser extent than those with an FEP. Moreover, the FEP group demonstrated more pronounced negative deviations, particularly in the hippocampus, suggesting a potential reduction in covariance between the hippocampus and other brain regions (Supplementary Figure 2).

**Figure 2.**
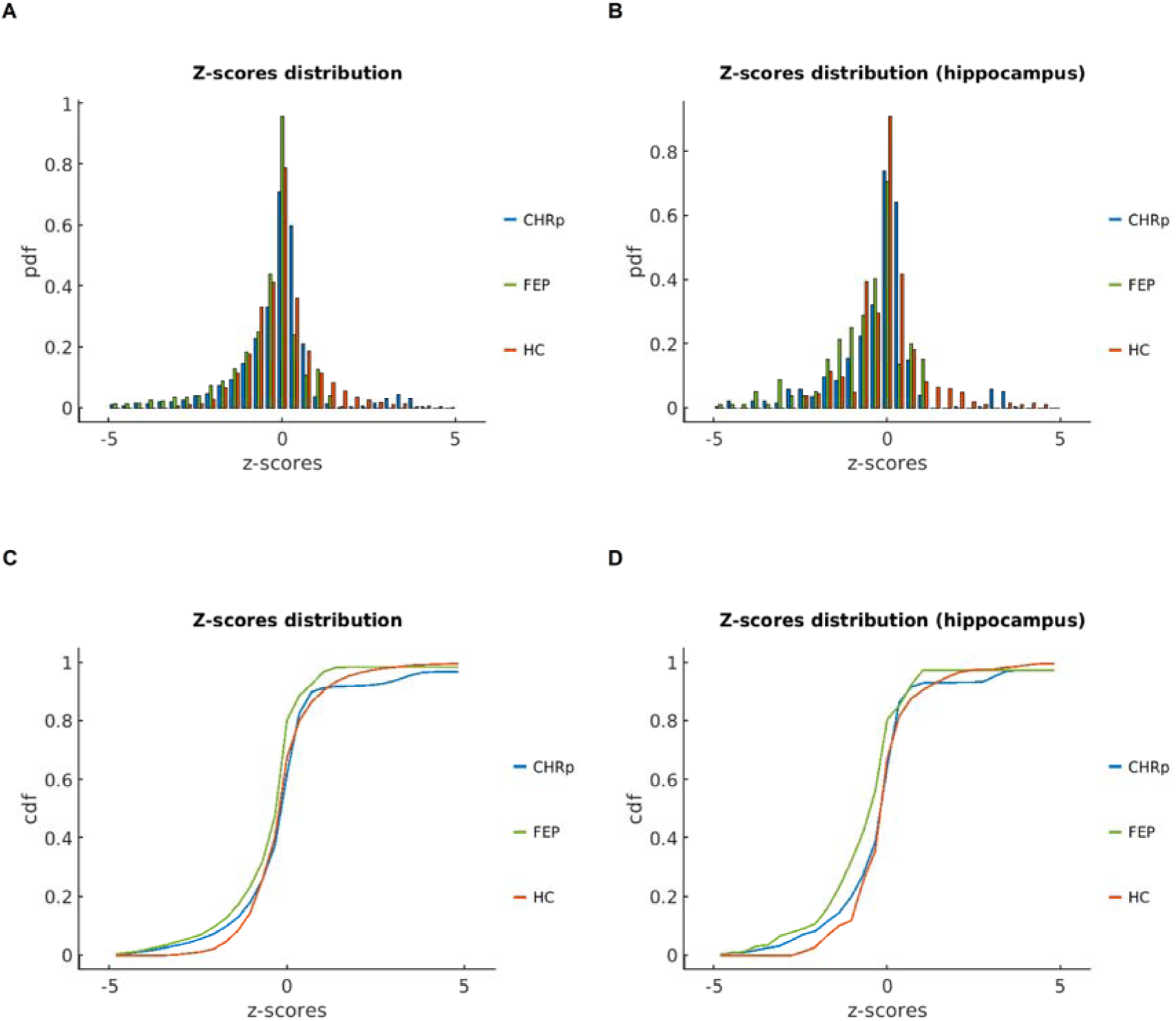
Perturbation covariance z-score statistics. Probability density function (PDF) of z-score distributions between healthy controls (HC), individuals at clinical high-risk for psychosis (CHRp) and people with a first episode of psychosis (FEP), across the whole brain (**A**) and focusing on correlations with the hippocampus only (**B**). Cumulative distribution function (CDF) of z-scores from the lower triangular matrices for HC, CHRp and FEP across the whole brain (**C**) and focusing specifically on the hippocampus (**D**).

## Discussion

The present study found that while hippocampal GABA_A_Rα5 availability did not differ between groups, brain-wide and hippocampal GABA_A_Rα5 covariance patterns were decreased in both CHRp and FEP individuals compared to HC. These findings suggest that psychosis symptoms may be associated with distributed rather than focal alterations in the GABA_A_Rα5 system.

Our findings build upon previous research investigating GABAergic function in psychosis, both at the microcircuit and whole-brain levels. Initial evidence for a GABAergic dysfunction in psychosis came from post-mortem studies in schizophrenia. These studies showed non-pyramidal cell reductions in the hippocampus [72], specifically the parvalbumin-positive (PVALB+) inhibitory interneurons [2,3]. Similarly, a disruption in hippocampal development in the MAM model led to local Pvalb+ interneuron loss [14,15], GABA_A_Rα5 density decrease across the hippocampus [25], subcortical hyperdopaminergia and behavioural changes of relevance for schizophrenia [14,16]. These psychosis-relevant phenotypes in MAM-treated rats were prevented by peripubertal benzodiazepine administration, and normalised by direct infusion of a GABA_A_Rα5 positive allosteric modulator into the hippocampus [21,22,24]. The present study sought to forward-translate these findings by investigating GABA_A_Rα5 availability in individuals at CHRp and with an FEP.

Contrary to our hypothesis, we found no evidence of alterations in [^11^C]Ro15-4513 binding in CHRp or FEP individuals compared to HC, either in the hippocampus or across the brain. To our knowledge, this is the first [^11^C]Ro15-4513 study in the CHRp population. The CHRp group was well-powered, making it unlikely for sample size to explain the null finding. These results may suggest that systematic alterations in regional GABA_A_Rα5 availability are not present in CHRp individuals. However, regional GABA_A_Rα5 deficits might only be present in a subgroup of CHRp individuals who subsequently develop psychosis. Although our sample was not large enough to investigate this, two previous studies addressed this question using less selective compounds. A recent study showed that normative GABA_A_R availability was not associated with brain perfusion differences between CHRp individuals with different transition status [73]. Additionally, a retrospective analysis in over 500 CHRp individuals found no association between benzodiazepine exposure and psychosis transition risk [74]. While this may suggest that general GABAergic function might not be a predictor of transition, future longitudinal studies using compounds specific to individual GABAergic system components are needed to elucidate this.

While we found no evidence of a difference in [^11^C]Ro15-4513 binding in the hippocampus of individuals with an FEP compared to HC, one previous [^11^C]Ro15-4513 study reported lower regional GABA_A_Rα5 availability in the hippocampus of ten antipsychotic-free patients with schizophrenia [38]. In contrast, another previous study with eleven participants (both medicated and medication-free) did not find such a difference [37]. As all studies to date have involved small numbers of participants, further investigation in larger samples is warranted. Nevertheless, it is plausible that changes in regional GABA_A_Rα5 availability may not be present in patients currently on antipsychotic medication. This is partly consistent with preclinical evidence showing increases in GABA_A_Rα5 availability following antipsychotic exposure [75], and a proton magnetic resonance spectroscopy (^1^H-MRS) study which reported normalised GABA levels in the anterior cingulate cortex of medication-naïve schizophrenia patients after antipsychotic treatment [76]. Further research directly comparing larger participant samples with and without antipsychotic treatment is required to expand on these findings. Such investigations in human participants *in vivo* would also circumvent limitations of post-mortem studies, such as disorder chronicity and effects of death.

Reviewing the common GABAergic interneuron-receptor subtype associations may also help understand the findings from port-mortem examinations in schizophrenia, which provided the central evidence towards GABAergic dysfunction in psychosis. PVALB+ inhibitory interneurons most commonly synapse onto α1 subunit-containing GABA_A_Rs, whereas GABA_A_Rα5s are expressed in proximity to somatostatin (SST)-expressing interneurons [23,77–79]. Previous research showed that about 66% of [^11^C]Ro15-4513 signal represents GABA_A_Rα5 binding [80]. In our previous imaging transcriptomics study, we found that [^11^C]Ro15-4513 binding tracked the expression of *GABRA5* and *SST*, and covaried negatively with *PVALB* expression [32]. This may suggest that [^11^C]Ro15-4513 PET, tracking GABA_A_Rα5s and SST+ inhibitory interneurons more likely than PVALB+ interneurons, may not be optimal for capturing alterations in the GABAergic microcircuits posited to be predominantly involved in psychosis symptom emergence by port-mortem examinations. However, it is plausible that a loss of PVALB+ interneurons may lead to regional GABA_A_Rα5 upregulation to compensate for a decrease in inhibition. Alternatively, any alterations in GABA_A_Rα5 expression may be compensated by changes in the levels of other GABA_A_Rs that [^11^C]Ro15-4513 binds to. Further research using novel radiotracers and other techniques such as *in vitro* cultures of interneurons is required to elucidate this question.

Our second hypothesis was that CHRp and FEP individuals would exhibit significant deviations in GABA_A_Rα5 network covariance relative to the normative healthy control group, both brain-wide and when focusing on the hippocampus, consistent with the notion that the hippocampus may play a key role in psychosis [81,82]. Accordingly, we identified significant covariance decrease in GABA_A_Rα5 availability in CHRp and FEP compared to HC individuals across brain regions, which was also present for the hippocampus. This was indicated by an average decrease in covariance after the addition of a participant from either of these groups to the reference group. These results are broadly consistent with previous studies on brain functional and structural variability in schizophrenia. For example, a recent study employing a normative modelling approach found increased variability in brain structure in schizophrenia [83]. At the temporal scale, there is robust meta-analytic evidence for altered resting-state functional connectivity across the psychosis spectrum [e.g., 84–86]. Moreover, several meta-analyses have found alterations in the brain-wide pattern of regional hyper- and hypoactivations in schizophrenia [87–89]. Overall, these results suggest that alterations in the organisation of the GABA_A_Rα5 system, both across the brain and between the hippocampus and other brain regions, may be involved in clinical high-risk and early psychosis, which aligns with the dysconnectivity hypothesis of schizophrenia [41]. It is plausible that this dysregulation contributes to the previously observed hippocampal hyperactivity. However, it is less well understood what molecular mechanisms may underlie such phenomenon. Future research should address this by investigating the relationships between GABA_A_Rα5 system organisation and other biological processes, such as glutamatergic neurotransmission.

There are several strengths of this study. Importantly, this is the first study investigating GABA_A_Rα5 availability in people at CHRp, for which we reached a sample size indicated by our power calculation. Additionally, we employed full tracer kinetic quantification and an additional molecular covariance approach to comprehensively characterise brain-wide GABA_A_Rα5 availability. Nevertheless, we did not reach the participant number indicated by power calculation in the FEP group, limiting the interpretability of these results. Further, we cannot exclude secondary binding to other targets by the radiotracer we employed. Pharmacological competition studies showed that even in regions rich in GABA_A_Rα5 expression, this receptor subtype accounts for 60-70% of all local [^11^C]Ro15-4513 binding [80]. Additionally, our [^11^C]Ro15-4513 binding covariance perturbation analyses relied on comparisons with the HC group using metrics derived from the HC group. As such, our results need to be corroborated by a similar study comparing a HC group alongside a CHRp and FEP sample with an independent reference HC group. Finally, we were not able to investigate if binding was related to clinical outcomes in CHRp individuals (such as transition to psychosis) because the sample size was too small.

Our findings suggest that individuals at CHRp and with an FEP show lower brain-wide GABA_A_Rα5 system covariance, but no regional availability alterations in the hippocampus. These findings support the utility of constructing individualised metabolic abnormality networks derived from single participant scans in uncovering molecular mechanisms underlying psychosis symptom severity.

## Acknowledgements

This research was funded in whole, or in part, by the Wellcome Trust & The Royal Society [Sir Henry Dale Fellowship 202397/Z/16/Z to GM]. PBL was in receipt of a PhD studentship funded by the National Institute for Health and Care Research (NIHR) Maudsley Biomedical Research Centre (BRC). The views expressed are those of the author and not necessarily those of the NHS, the NIHR or the Department of Health and Social Care.

AAG received funding from USPHS NIMH MH57440. MV is supported by EU funding within the MUR PNRR “National Center for HPC, BIG DATA AND QUANTUM COMPUTING (Project no. CN00000013 CN1), the Ministry of University and Research within the Complementary National Plan PNC DIGITAL LIFELONG PREVENTION - DARE (Project no PNC0000002_DARE), and by Fondo per il Programma Nazionale di Ricerca e Progetti di Rilevante Interesse Nazionale (PRIN, Project no 2022RXM3H7).

The authors would like to thank Dr Zerrin Atakan for providing medical expertise during participant recruitment.

For the purposes of open access, the author has applied a Creative Commons Attribution (CC BY) licence to any Accepted Author Manuscript version arising from this submission.

## Data Availability

The positron emission tomography data will be made publicly available upon publication. Any other data will be available from the corresponding author on reasonable request.

