## Supplement for "GABA_A_ receptor availability in clinical high-risk and first-episode psychosis: a [^11^C]Ro15-4513 positron emission tomography study"

[2.	Supplementary Results: Hippocampal [^11^C]Ro15-4513 binding	3](#_Toc191381918)

[3.	Supplementary Results: Brain-wide [^11^C]Ro15-4513 binding	3](#_Toc191381919)

[6.	Supplementary Methods and Results: Group classification by regional [^11^C]Ro15-4513 binding covariance	13](#_Toc191381922)

### 1. Supplementary Results: Demographic and clinical data

Pairwise comparison of age between study groups revealed that the FEP participants were older than the other two groups, although the results did not survive multiple comparison correction (Supplementary Table 1). Additionally, CHRp participants reported more symptoms of anxiety and depression than FEP and HC participants, and FEP participants reported more symptoms of anxiety and depression than HC participants (Supplementary Table 1).

|  | Comparison | n1 | n2 | p | p.signif | p.adj | p.adj.signif |
| --- | --- | --- | --- | --- | --- | --- | --- |
| Age | CHRp-Control | 22 | 23 | 0.988 | ns | 0.988 | ns |
|  | CHRp-FEP | 22 | 10 | 0.023 | * | 0.0646 | ns |
|  | Control-FEP | 23 | 10 | 0.0215 | * | 0.0646 | ns |
| HAM-A | CHRp-Control | 22 | 23 | 9.92E-08 | **** | 2.98E-07 | **** |
|  | CHRp-FEP | 22 | 10 | 0.0181 | * | 0.0304 | * |
|  | Control-FEP | 23 | 10 | 0.0152 | * | 0.0304 | * |
| HAM-D | CHRp-Control | 22 | 23 | 6.38E-09 | **** | 1.91E-08 | **** |
|  | CHRp-FEP | 22 | 10 | 0.0464 | * | 0.0464 | * |
|  | Control-FEP | 23 | 10 | 0.000938 | *** | 0.00188 | ** |
| GF:S | CHRp-Control | 22 | 23 | 0.00144 | ** | 0.00288 | ** |
|  | CHRp-FEP | 22 | 10 | 0.351 | ns | 0.351 | ns |
|  | Control-FEP | 23 | 10 | 0.000701 | *** | 0.0021 | ** |
| GF:R | CHRp-Control | 22 | 23 | 0.00086 | *** | 0.00172 | ** |
|  | CHRp-FEP | 22 | 10 | 0.172 | ns | 0.172 | ns |
|  | Control-FEP | 23 | 10 | 0.000113 | *** | 0.00034 | *** |

**Supplementary Table 1. Pairwise comparisons of age and reported anxiety and depression in the study samples.** CHRp, clinical high risk for psychosis; FEP, First Episode of Psychosis; GF:S/R, Global Functioning: Social/Role Scale; HAM-A/D, Hamilton Anxiety/Depression Scale; ns, not significant; p.signif, p-value significance; p.adj, adjusted p-value, p.adj.signif, adjusted p-value significance.

### 2. Supplementary Results: Hippocampal [^11^C]Ro15-4513 binding

#### 2.1. Hippocampal [^11^C]Ro15-4513 binding: CHRp versus HC

As a sensitivity check, we performed the one-way ANOVA including hippocampal [^11^C]Ro15-4513 binding values just between the groups which numbers met our power calculation sample size, namely the CHRp and HC groups. There was no significant difference in this analysis (one-way ANOVA with sex and age as covariates of no interest: F(1,41)=0.33, p=0.57, ges=0.008).

#### 2.2. Hippocampal [^11^C]Ro15-4513 binding and symptoms

In HC, hippocampal [^11^C]Ro15-4513 binding correlated positively with CAARMS positive symptoms (Pearson’s r=0.57, p=0.005), CAARMS negative symptoms (Pearson’s r=0.64, p=0.02) and HAM-A anxiety symptoms (Pearson’s r=0.64, p=0.002), but not with HAM-D depression symptoms (Pearson’s r=0.08, p=0.73). In individuals at CHRp, hippocampal [^11^C]Ro15-4513 binding did not correlate with either symptom dimension (CAARMS-positive: Pearson’s r=-0.33, p=0.13; CAARMS-negative: Pearson’s r=0.18, p=0.43; HAM-A: Pearson’s r=0.27, p=0.23; HAM-D: Pearson’s r=0.24, p=0.28). In people with an FEP, hippocampal [^11^C]Ro15-4513 binding correlated with PANSS-negative (Pearson’s r=-0.65, p=0.04) but not with other symptoms (PANSS-positive: Pearson’s r=0.1, p=0.78; HAM-A: Pearson’s r=0.35, p=0.32; HAM-D: Pearson’s r=-0.1, p=0.78).

### 3. Supplementary Results: Brain-wide [^11^C]Ro15-4513 binding

|  | [^11^C]Ro15-4513 V_T_ mean (SD) | | |
| --- | --- | --- | --- |
|  | HC | CHRp | FEP |
| Brain | 4.94 (0.72) | 4.75 (0.66) | 4.70 (0.61) |
| Grey Matter | 5.42 (0.79) | 5.20 (0.71) | 5.16 (0.69) |
| Cortical | 5.97 (0.89) | 5.71 (0.79) | 5.67 (0.75) |
| Subcortical | 4.15 (0.6) | 4.15 (0.69) | 4.14 (0.47) |
| Cerebellum | 3.00 (0.37) | 2.93 (0.45) | 2.89 (0.43) |
| Brain Stem | 1.27 (0.15) | 1.29 (0.34) | 1.19 (0.15) |
| Occipital Lobe | 5.15 (0.76) | 5.01 (0.71) | 5.06 (0.69) |
| Insular Cortex | 10.86 (1.67) | 10.51 (1.57) | 10.51 (1.37) |
| Temporal Lobe | 7.07 (1.06) | 6.82 (1) | 6.64 (0.77) |
| Frontal Lobe | 5.49 (0.86) | 5.23 (0.74) | 5.19 (0.78) |
| Cingulate Cortex | 8.56 (1.33) | 8.29 (1.29) | 8.10 (0.96) |
| Parietal Lobe | 5.27 (0.8) | 4.99 (0.68) | 5.05 (0.68) |
| Basal Ganglia | 5.10 (0.8) | 5.05 (0.93) | 5.17 (0.64) |
| Thalamus | 3.25 (0.45) | 3.31 (0.55) | 3.18 (0.41) |
| Parahippocampal Gyrus | 8.67 (3.13) | 7.88 (1.06) | 7.66 (1.13) |
| Amygdala | 7.80 (1.25) | 7.66 (1.3) | 7.88 (1.09) |
| Hippocampus | 9.64 (1.44) | 9.42 (1.54) | 9.14 (1.36) |
| Anterior Cingulate | 26.04 (79.69) | 9.08 (1.42) | 8.90 (1.06) |
| Globus Pallidus | 2.32 (0.31) | 2.29 (0.48) | 2.34 (0.24) |
| Striatum | 5.60 (0.9) | 5.54 (1.03) | 5.66 (0.7) |
| Caudate | 4.39 (0.76) | 4.39 (0.92) | 4.34 (0.72) |
| Nucleus Accumbens | 65.25 (255.24) | 12.93 (6.18) | 11.87 (1.42) |
| Putamen | 4.87 (0.78) | 4.82 (0.9) | 5.07 (0.63) |

**Supplementary Table 2. Regional [^11^C]Ro15-4513 binding in the healthy control (HC), clinical high-risk for psychosis (CHRp) and first episode of psychosis (FEP) groups.** SD, standard deviation; V_T_, total volume of distribution.

##### 3.1. Brain-wide comparisons: CHRp vs HC

| Region | Variable | DFn | DFd | F | p(uncorr) | p<.05 | ges |
| --- | --- | --- | --- | --- | --- | --- | --- |
| Brain | Group | 1 | 28 | 0.135 | 0.716 |  | 0.005 |
|  | Age | 1 | 28 | 0.484 | 0.492 |  | 0.017 |
|  | Sex | 1 | 28 | 0.845 | 0.366 |  | 0.029 |
| Grey matter | Group | 1 | 28 | 0.136 | 0.715 |  | 0.005 |
|  | Age | 1 | 28 | 0.448 | 0.509 |  | 0.016 |
|  | Sex | 1 | 28 | 0.498 | 0.486 |  | 0.017 |
| Cortical | Group | 1 | 28 | 0.148 | 0.704 |  | 0.005 |
|  | Age | 1 | 28 | 0.67 | 0.42 |  | 0.023 |
|  | Sex | 1 | 28 | 0.368 | 0.549 |  | 0.013 |
| Subcortical | Group | 1 | 28 | 0.046 | 0.831 |  | 0.002 |
|  | Age | 1 | 28 | 0.376 | 0.545 |  | 0.013 |
|  | Sex | 1 | 28 | 2.295 | 0.141 |  | 0.076 |
| Cerebellum | Group | 1 | 28 | 0.002 | 0.962 |  | 8.37E-05 |
|  | Age | 1 | 28 | 0.424 | 0.52 |  | 0.015 |
|  | Sex | 1 | 28 | 1.713 | 0.201 |  | 0.058 |
| Brain stem | Group | 1 | 28 | 0.626 | 0.436 |  | 0.022 |
|  | Age | 1 | 28 | 1.844 | 0.185 |  | 0.062 |
|  | Sex | 1 | 28 | 2.289 | 0.142 |  | 0.076 |
| Occipital lobe | Group | 1 | 28 | 0.26 | 0.614 |  | 0.009 |
|  | Age | 1 | 28 | 0.145 | 0.706 |  | 0.005 |
|  | Sex | 1 | 28 | 0.422 | 0.521 |  | 0.015 |
| Insular cortex | Group | 1 | 28 | 0.237 | 0.63 |  | 0.008 |
|  | Age | 1 | 28 | 0.11 | 0.742 |  | 0.004 |
|  | Sex | 1 | 28 | 1.304 | 0.263 |  | 0.044 |
| Temporal lobe | Group | 1 | 28 | 0.014 | 0.907 |  | 0.000499 |
|  | Age | 1 | 28 | 0.321 | 0.576 |  | 0.011 |
|  | Sex | 1 | 28 | 0.919 | 0.346 |  | 0.032 |
| Frontal lobe | Group | 1 | 28 | 0.148 | 0.704 |  | 0.005 |
|  | Age | 1 | 28 | 1.125 | 0.298 |  | 0.039 |
|  | Sex | 1 | 28 | 0.09 | 0.766 |  | 0.003 |
| Cingulate cortex | Group | 1 | 28 | 0.137 | 0.714 |  | 0.005 |
|  | Age | 1 | 28 | 0.558 | 0.461 |  | 0.02 |
|  | Sex | 1 | 28 | 1.72 | 0.2 |  | 0.058 |
| Parietal lobe | Group | 1 | 28 | 0.285 | 0.598 |  | 0.01 |
|  | Age | 1 | 28 | 0.555 | 0.463 |  | 0.019 |
|  | Sex | 1 | 28 | 0.038 | 0.847 |  | 0.001 |
| Basal ganglia | Group | 1 | 28 | 0.171 | 0.682 |  | 0.006 |
|  | Age | 1 | 28 | 0.58 | 0.453 |  | 0.02 |
|  | Sex | 1 | 28 | 1.434 | 0.241 |  | 0.049 |
| Thalamus | Group | 1 | 28 | 0.122 | 0.729 |  | 0.004 |
|  | Age | 1 | 28 | 0.312 | 0.581 |  | 0.011 |
|  | Sex | 1 | 28 | 1.89 | 0.18 |  | 0.063 |
| Parahippocampal gyrus | Group | 1 | 28 | 0.019 | 0.892 |  | 0.000672 |
|  | Age | 1 | 28 | 0.165 | 0.688 |  | 0.006 |
|  | Sex | 1 | 28 | 2.131 | 0.155 |  | 0.071 |
| Amygdala | Group | 1 | 28 | 0.434 | 0.515 |  | 0.015 |
|  | Age | 1 | 28 | 0.027 | 0.87 |  | 0.000966 |
|  | Sex | 1 | 28 | 0.379 | 0.543 |  | 0.013 |
| Hippocampus | Group | 1 | 28 | 0.000522 | 0.982 |  | 1.86E-05 |
|  | Age | 1 | 28 | 0.228 | 0.637 |  | 0.008 |
|  | Sex | 1 | 28 | 0.655 | 0.425 |  | 0.023 |
| Anterior cingulate | Group | 1 | 28 | 0.133 | 0.718 |  | 0.005 |
|  | Age | 1 | 28 | 0.52 | 0.477 |  | 0.018 |
|  | Sex | 1 | 28 | 1.481 | 0.234 |  | 0.05 |
| Globus pallidus | Group | 1 | 28 | 0.099 | 0.756 |  | 0.004 |
|  | Age | 1 | 28 | 2.113 | 0.157 |  | 0.07 |
|  | Sex | 1 | 28 | 2.814 | 0.105 |  | 0.091 |
| Striatum | Group | 1 | 28 | 0.14 | 0.711 |  | 0.005 |
|  | Age | 1 | 28 | 0.535 | 0.471 |  | 0.019 |
|  | Sex | 1 | 28 | 1.172 | 0.288 |  | 0.04 |
| Caudate | Group | 1 | 28 | 0.016 | 0.9 |  | 0.000578 |
|  | Age | 1 | 28 | 0.354 | 0.556 |  | 0.013 |
|  | Sex | 1 | 28 | 0.454 | 0.506 |  | 0.016 |
| Nucleus accumbens | Group | 1 | 28 | 1.829 | 0.187 |  | 0.061 |
|  | Age | 1 | 28 | 2.094 | 0.159 |  | 0.07 |
|  | Sex | 1 | 28 | 1.093 | 0.305 |  | 0.038 |
| Putamen | Group | 1 | 28 | 0.441 | 0.512 |  | 0.016 |
|  | Age | 1 | 28 | 1.431 | 0.242 |  | 0.049 |
|  | Sex | 1 | 28 | 1.744 | 0.197 |  | 0.059 |

**Supplementary Table 3. Post-hoc pairwise comparisons between the healthy control and clinical high risk for psychosis groups within all regions tested.** ges, generalised eta squared (effect size).

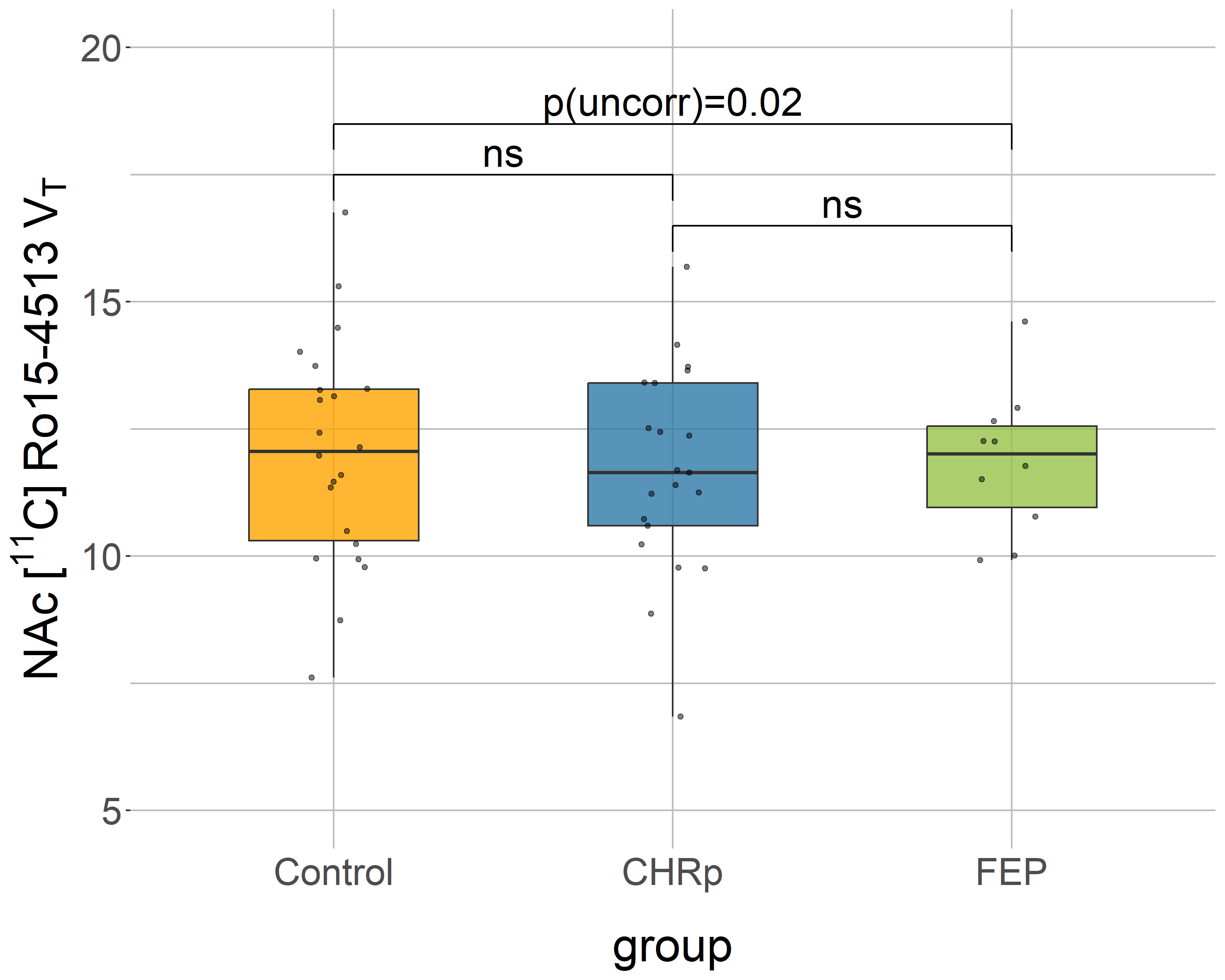

**Supplementary Figure 1. GABA_A_Rα5 availability in the nucleus accumbens (NAc) between groups**. Lower NAc GABA_A_Rα5 availability in first-episode of psychosis (FEP) compared to healthy controls (HC) at the uncorrected level. CHRp, clinical high risk for psychosis; FEP, first episode of psychosis; ns, not significant; V_T_, total volume of distribution. Two data points removed for illustrative purposes.

##### 3.2. Brain-wide comparisons: FEP vs HC

| Region | Variable | DFn | DFd | F | P(uncorr) | p<.05 | ges |
| --- | --- | --- | --- | --- | --- | --- | --- |
| Brain | Group | 1 | 29 | 0.511 | 0.48 |  | 0.017 |
|  | Age | 1 | 29 | 1.909 | 0.178 |  | 0.062 |
|  | Sex | 1 | 29 | 2.497 | 0.125 |  | 0.079 |
| Grey matter | Group | 1 | 29 | 0.523 | 0.475 |  | 0.018 |
|  | Age | 1 | 29 | 1.677 | 0.205 |  | 0.055 |
|  | Sex | 1 | 29 | 2.746 | 0.108 |  | 0.086 |
| Cortical | Group | 1 | 29 | 0.557 | 0.461 |  | 0.019 |
|  | Age | 1 | 29 | 1.867 | 0.182 |  | 0.06 |
|  | Sex | 1 | 29 | 2.882 | 0.1 |  | 0.09 |
| Subcortical | Group | 1 | 29 | 0.002 | 0.966 |  | 6.30E-05 |
|  | Age | 1 | 29 | 0.565 | 0.458 |  | 0.019 |
|  | Sex | 1 | 29 | 1.284 | 0.266 |  | 0.042 |
| Cerebellum | Group | 1 | 29 | 0.586 | 0.45 |  | 0.02 |
|  | Age | 1 | 29 | 0.301 | 0.587 |  | 0.01 |
|  | Sex | 1 | 29 | 1.45 | 0.238 |  | 0.048 |
| Brain stem | Group | 1 | 29 | 1.179 | 0.286 |  | 0.039 |
|  | Age | 1 | 29 | 0.93 | 0.343 |  | 0.031 |
|  | Sex | 1 | 29 | 0.828 | 0.37 |  | 0.028 |
| Occipital lobe | Group | 1 | 29 | 0.178 | 0.677 |  | 0.006 |
|  | Age | 1 | 29 | 0.436 | 0.514 |  | 0.015 |
|  | Sex | 1 | 29 | 2.323 | 0.138 |  | 0.074 |
| Insular cortex | Group | 1 | 29 | 0.392 | 0.536 |  | 0.013 |
|  | Age | 1 | 29 | 0.167 | 0.686 |  | 0.006 |
|  | Sex | 1 | 29 | 1.04 | 0.316 |  | 0.035 |
| Temporal lobe | Group | 1 | 29 | 1.111 | 0.301 |  | 0.037 |
|  | Age | 1 | 29 | 0.359 | 0.554 |  | 0.012 |
|  | Sex | 1 | 29 | 1.199 | 0.282 |  | 0.04 |
| Frontal lobe | Group | 1 | 29 | 0.494 | 0.488 |  | 0.017 |
|  | Age | 1 | 29 | 3.881 | 0.058 |  | 0.118 |
|  | Sex | 1 | 29 | 5.272 | 0.029 | * | 0.154 |
| Cingulate cortex | Group | 1 | 29 | 0.349 | 0.559 |  | 0.012 |
|  | Age | 1 | 29 | 2.05 | 0.163 |  | 0.066 |
|  | Sex | 1 | 29 | 1.305 | 0.263 |  | 0.043 |
| Parietal lobe | Group | 1 | 29 | 0.303 | 0.586 |  | 0.01 |
|  | Age | 1 | 29 | 1.961 | 0.172 |  | 0.063 |
|  | Sex | 1 | 29 | 2.633 | 0.115 |  | 0.083 |
| Basal ganglia | Group | 1 | 29 | 0.007 | 0.932 |  | 0.000252 |
|  | Age | 1 | 29 | 0.002 | 0.966 |  | 6.46E-05 |
|  | Sex | 1 | 29 | 1.219 | 0.279 |  | 0.04 |
| Thalamus | Group | 1 | 29 | 0.016 | 0.899 |  | 0.000562 |
|  | Age | 1 | 29 | 1.636 | 0.211 |  | 0.053 |
|  | Sex | 1 | 29 | 0.837 | 0.368 |  | 0.028 |
| Parahippocampal gyrus | Group | 1 | 29 | 2.723 | 0.11 |  | 0.086 |
|  | Age | 1 | 29 | 0.853 | 0.363 |  | 0.029 |
|  | Sex | 1 | 29 | 1.613 | 0.214 |  | 0.053 |
| Amygdala | Group | 1 | 29 | 0.027 | 0.871 |  | 0.000925 |
|  | Age | 1 | 29 | 0.101 | 0.753 |  | 0.003 |
|  | Sex | 1 | 29 | 2.737 | 0.109 |  | 0.086 |
| Hippocampus | Group | 1 | 29 | 0.721 | 0.403 |  | 0.024 |
|  | Age | 1 | 29 | 0.069 | 0.794 |  | 0.002 |
|  | Sex | 1 | 29 | 0.251 | 0.62 |  | 0.009 |
| Anterior cingulate | Group | 1 | 29 | 0.504 | 0.483 |  | 0.017 |
|  | Age | 1 | 29 | 0.83 | 0.37 |  | 0.028 |
|  | Sex | 1 | 29 | 1.006 | 0.324 |  | 0.034 |
| Globus pallidus | Group | 1 | 29 | 0.019 | 0.892 |  | 0.000649 |
|  | Age | 1 | 29 | 0.033 | 0.858 |  | 0.001 |
|  | Sex | 1 | 29 | 0.152 | 0.7 |  | 0.005 |
| Striatum | Group | 1 | 29 | 0.019 | 0.891 |  | 0.00066 |
|  | Age | 1 | 29 | 0.000661 | 0.98 |  | 2.28E-05 |
|  | Sex | 1 | 29 | 1.295 | 0.265 |  | 0.043 |
| Caudate | Group | 1 | 29 | 0.151 | 0.7 |  | 0.005 |
|  | Age | 1 | 29 | 0.176 | 0.678 |  | 0.006 |
|  | Sex | 1 | 29 | 2.383 | 0.134 |  | 0.076 |
| Nucleus accumbens | Group | 1 | 29 | 6.047 | 0.02 | * | 0.173 |
|  | Age | 1 | 29 | 14.445 | 0.000685 | * | 0.332 |
|  | Sex | 1 | 29 | 0.22 | 0.643 |  | 0.008 |
| Putamen | Group | 1 | 29 | 0.057 | 0.814 |  | 0.002 |
|  | Age | 1 | 29 | 0.017 | 0.898 |  | 0.000577 |
|  | Sex | 1 | 29 | 1.586 | 0.218 |  | 0.052 |

**Supplementary Table 4.** **Post-hoc pairwise comparisons between the healthy control and first episode of psychosis groups within all regions tested.** ges, generalised eta squared (effect size).

### 4. Supplementary Results: Covariance perturbation analyses

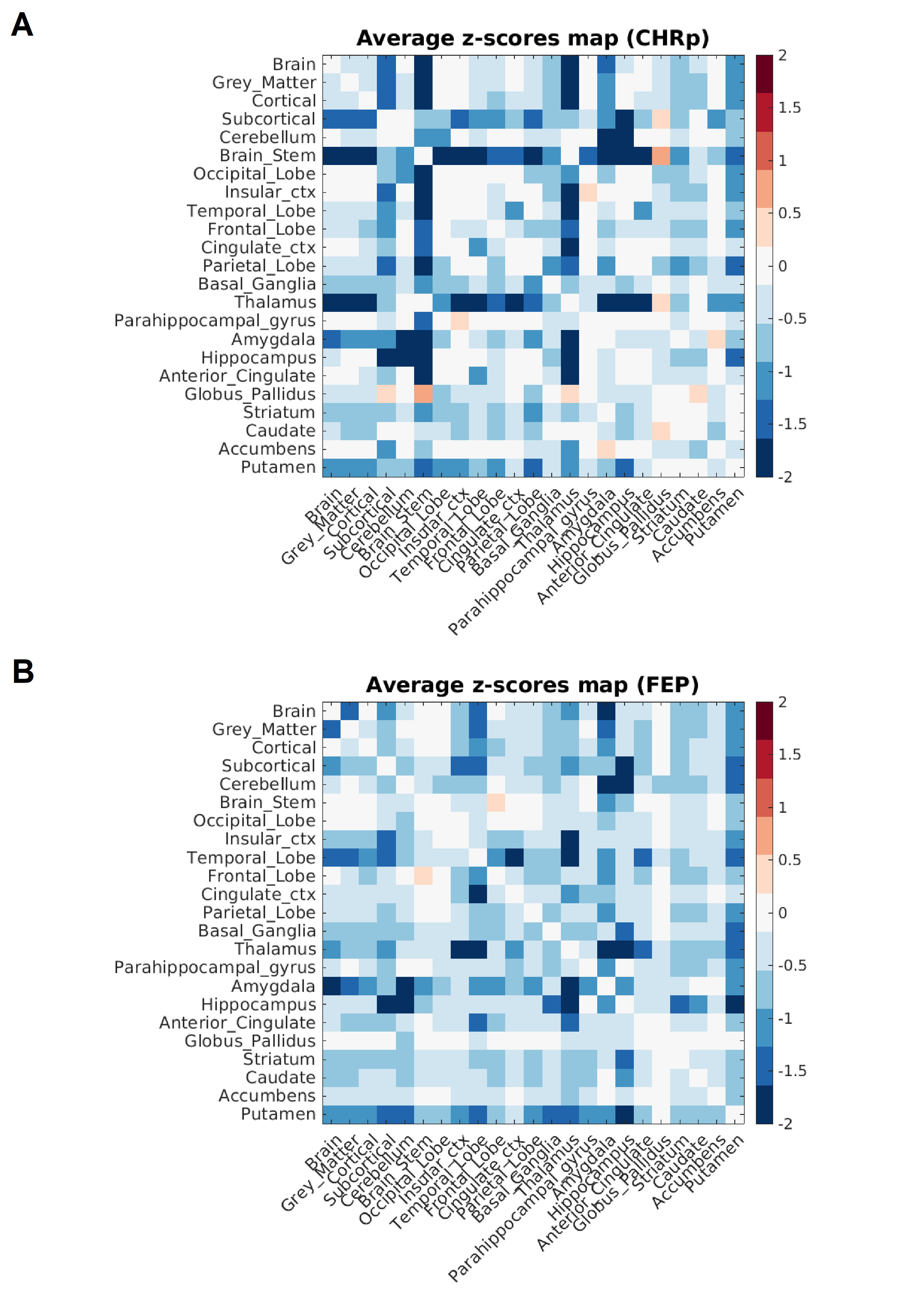

**Supplementary Figure 2.** **Average z-score matrix across participants for the clinical study groups.** Matrix for clinical high risk for psychosis (CHRp) (**A**), and for the first episode of psychosis (FEP) (**B**).

### 5. Supplementary Methods and Results: Regional extreme deviations analysis

###### 5.1. Supplementary Methods: Regional extreme deviations analysis

From the single-participant matrices, extreme deviations can be extracted to characterize deviation patterns at region levels for each participant. This analysis begins with thresholding the matrices at |z| > 2, identifying z-scores (deviations of the matrix) that are deemed "extreme" relative to the reference control group. A z-score value of 1.96 corresponds to a significance level of 0.05. The matrices are then binarized to mark the presence or absence of these extreme deviations. Subsequently, the regional extreme deviations are extracted:

Let $A_{j}$ represent a binarized symmetric $m \times m$matrix of deviations for a given participant *j,* where $m$ are the number of regions*.* For each participant *j*, we want to compute the mean of the elements in each row of matrix $A_{j}$. The mean of the elements in the *i-th* row of matrix $A_{j}$ for participant *j* is given by:

$$\mu_{ij}= \frac{1}{m}\sum_{k=1}^{m} a_{ik}$$

Where $\mu_{ij}$ is the mean of the elements in the i-th row of matrix $A_{j}$ for participant j, $a_{ik}$ denotes the element in the i-th row and k-th column of matrix $A_{j}$, and m represents the number of regions. After calculating the mean for each row, we construct a mean vector $\mu_{j}$ for each participant j, where each element of the vector corresponds to the mean of a row in matrix $A_{j}$:

$$\mu_{j}= \left[ \begin{matrix} \mu_{j1} & \mu_{j2} \ldots\end{matrix} \begin{matrix} \mu_{jm-1} & \mu_{jm} \end{matrix} \right]$$

Ultimately, we obtain a matrix M of dimension $s\times m$, where each row $s$ corresponds to a participant, and each column $m$ corresponds to a region.

$$M= \left[ \begin{matrix} \mu_{11} & \cdots& \mu_{1m} \\ \vdots& \ddots& \vdots\\ \mu_{s1} & \cdots& \mu_{sm} \end{matrix} \right]$$

The element at position $\left( j,i \right)$ represents the mean deviation for region i in participant j. This regional metric is subsequently utilized to perform statistical comparisons of deviation patterns between groups.

Statistical comparisons of the regional extreme deviations were conducted using a 1,000-permutation test. The magnitude of the effect was quantified using Cohen's d effect size. These differences were first evaluated across all brain regions and subsequently focusing on the hippocampus.

Statistical correlations between the mean regional extreme deviations of the CHRp and FEP groups and clinical symptoms were assessed using Spearman's correlation. Specifically, the correlations between the mean regional extreme deviations in the CHRp group and positive symptom scores from the Comprehensive Assessment for At-Risk Mental States (CAARMS) [1], Hamilton Anxiety Rating Scale (HAM-A) [2], and Hamilton Depression Rating Scale (HAM-D) [3] were analysed. Similarly, correlations were calculated for the regional extreme deviations in the FEP group and positive symptom scores from the Positive and Negative Syndrome scale (PANSS) [4], HAM-A scores and HAM-D scores.

###### 5.2. Supplementary Results: Regional extreme deviations analysis

1,000-permutation tests revealed significant differences in regional extreme deviations when considering all regions between HC and FEP (p < 0.001, Cohen’s d = 1.16), between HC and CHRp (p < 0.0001, Cohen’s d = 3.05, and between FEP and CHRp (p < 0.001, Cohen’s d = 1.02). In contrast no statistical differences were observed between HC and FEP (p = 0.31, Cohen’s d = 0.38), HC and CHRp (p = 0.18, Cohen’s d = 0.39), and FEP and CHRp (p = 0.83, Cohen’s d = 0.10) when focusing exclusively on the extreme deviations on the hippocampus (Supplementary Figure 3). This could be due to the high variability in the deviations of hippocampus across the participants.

###### 5.3. Supplementary Results: Correlations of regional extreme deviations with symptoms

Correlation analysis yielded no statistically significant correlation between mean regional extreme deviations of CHRp with CAARMS positive scores (ρ = -0.15, p = 0.29), HAM-A scores (ρ = 0.25, p = 0.18), and HAM-D scores (ρ = 0.25, p = 0.15). Similarly, no statistically significant correlation was found between mean regional extreme deviation of FEP with PANSS positive scores (ρ = -0.30, p = 0.38), HAM-A scores (ρ = 0.49, p = 0.14), and HAM-D scores (ρ = 0.30, p = 0.38).

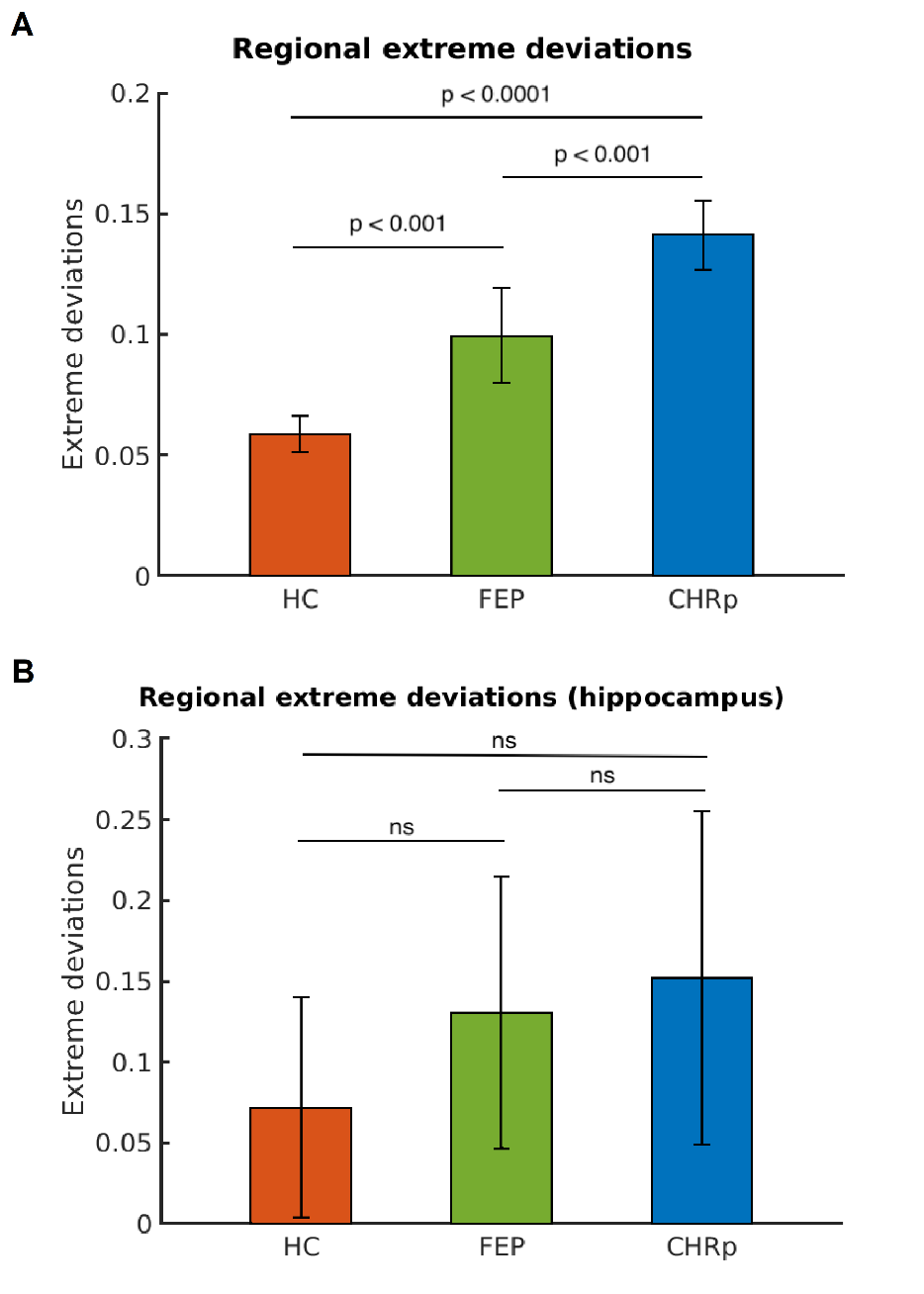

**Supplementary Figure 3. Average regional extreme deviations between healthy controls (HC), first episode of psychosis (FEP), and clinical high-risk for psychosis (CHRp).** Average extreme deviations considering all the brain regions (**A**) and considering only the hippocampus (**B**).

### 6. Supplementary Methods and Results: Group classification by regional [^11^C]Ro15-4513 binding covariance

###### 6.1. Supplementary Methods: Group classification by regional [^11^C]Ro15-4513 binding covariance

An additional data-driven analysis was conducted to assess whether the original study groups (HC, CHRp, and FEP) could be reclassified based solely on the deviations in [^11^C]Ro15-4513 correlation values. In this analysis, the vectorized lower triangular deviation matrices for each participant were employed as features in a Cubic Support Vector Machine (SVM) model configured in Matlab, utilizing 10-fold cross-validation (CV) with 10 repetitions, using an 80-20% split for the training and test sets, while accounting for class imbalance. The following hyperparameters were configured: regularization parameter C=3 and kernel type = cubic. The classification performance was evaluated using accuracy (ACC), area under the receiver operating characteristic curve (AUC), true positive rate (TPR), and true negative rate (TNR).

###### 6.2. Supplementary Results: Group classification by regional [^11^C]Ro15-4513 binding covariance

Classification analysis yielded moderate evidence towards significant between-group differences based on [^11^C]Ro15-4513 binding network deviations in covariance. Both classifications exceeded the chance-level accuracy (50%), with moderate differentiation achieved for both HC and CHRp (ACC = 59.7, AUC = 0.72, TPR = 0.34, TNR = 0.83) and FEP and HC (ACC = 69.3, AUC = 0.67, TPR = 0.30, TNR = 0.86). The low TPR values for both models indicated that the clinical groups were not easily classified by the models, suggesting differences may not have been consistent across all participants within the groups. The performance of both models was highly influenced by the split and assignment process, showing significant variability, as indicated by the large confidence intervals of the average ROC AUC.

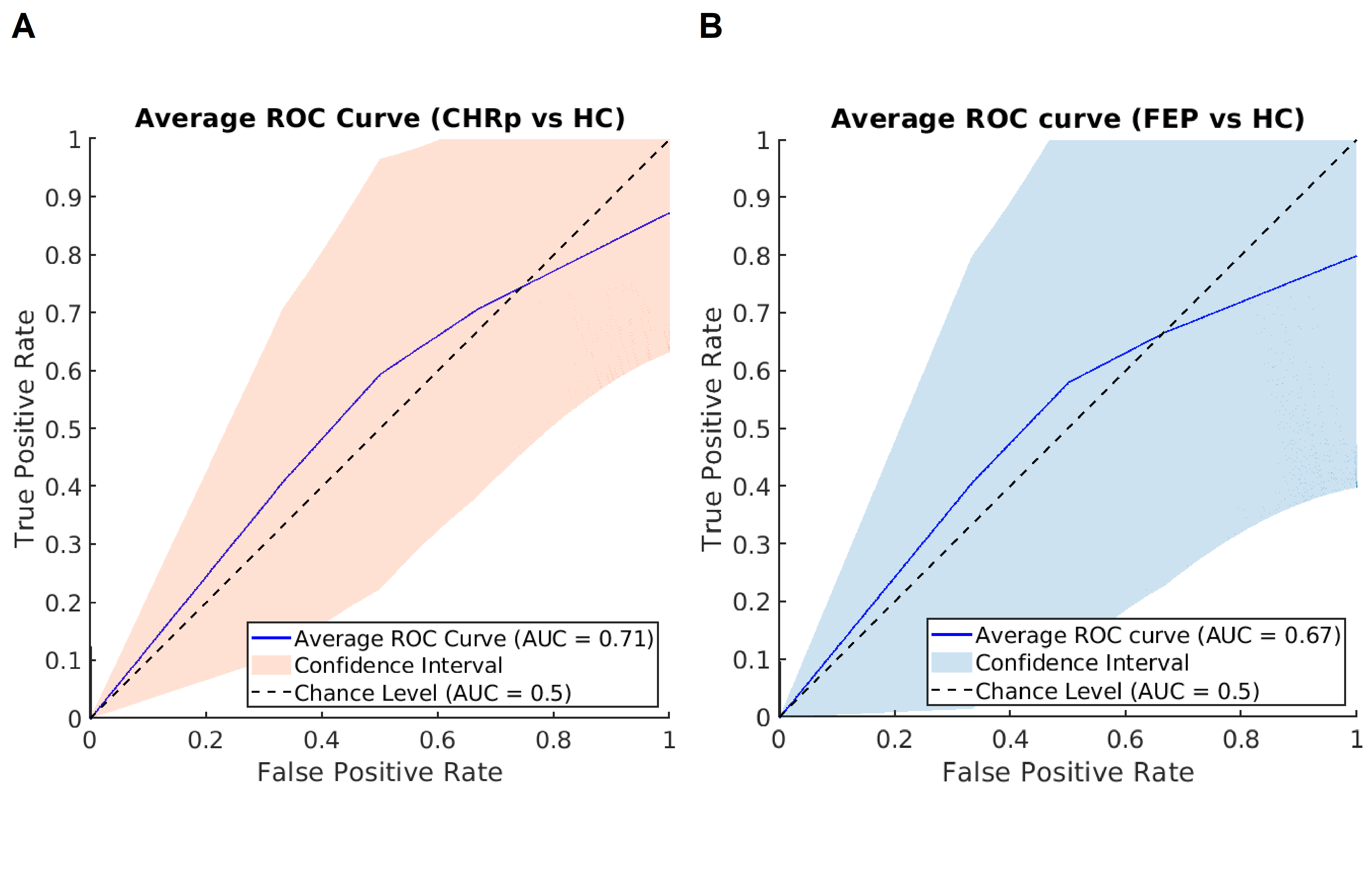
**Supplementary Figure 4. Group classification performance.** Moderate performance and high variability (AUC = 0.72, TPR = 0.34, TNR = 0.83) in the classification between healthy controls (HC) and individuals at clinical high-risk for psychosis (CHRp) (**A**). Moderate performance and high variability in the classification between HC and people with a first episode of psychosis (FEP) (AUC = 0.67, TPR = 0.3, TNR = 0.86) (**B**).
